# Cerebral glutamate levels over two years in initially antipsychotic-naïve first-episode patients with psychosis are related to clinical symptoms and cognition

**DOI:** 10.1101/2025.04.10.25325581

**Authors:** Kirsten Borup Bojesen, Cecilie Koldbæk Lemvigh, Anne Korning Sigvard, Mark Bitsch Vestergaard, Henrik Bo Wiberg Larsson, Egill Rostrup, Bjørn Hylsebeck Ebdrup, Birte Glenthøj

**Affiliations:** Center for Neuropsychiatric Schizophrenia Research (CNSR) & Center for Clinical Intervention and Neuropsychiatric Schizophrenia Research (CINS), Mental Health Center Glostrup, Copenhagen University Hospital – Mental Health Services CPH, Denmark; Functional Imaging Unit, Department of Clinical Physiology and Nuclear Medicine, Rigshospitalet Glostrup, University of Copenhagen, Denmark; Faculty of Health and Medical Sciences, Department of Clinical Medicine, University of Copenhagen, Denmark

**Author notes:** Corresponding author: CNSR, Mental Health Center Glostrup, Nordstjernevej 41, 2600 Glostrup, Denmark.

**Keywords:** Glutamate, ACC, thalamus, 1H-MRS, antipsychotic-naïve, first-episode, psychosis, schizophrenia

## Abstract

Although emerging evidence supports glutamatergic dysfunction in schizophrenia, clinical trials with glutamatergic compounds have overall been negative. This may be due to changes in glutamate levels during the course of illness.

To address this, we measured glutamate levels in dorsal anterior cingulate cortex (dACC) and left thalamus in 57 initially antipsychotic-naïve patients with first-episode psychosis (FEP) aged 22.6 ± 5.0 years (58% females) and 55 healthy controls (HC) on a 3T MR scanner at baseline, after six weeks (48 FEP and 53 HC), six months (37 FEP and 49 HC), and two years (35 FEP and 45 HC). Positive and negative symptoms and cognitive function in tests of attention and spatial working memory were assessed at all visits. Linear mixed models were used in statistical analyses.

We found lower glutamate levels in dACC in FEP (p=0.03) that was associated with deficits in attention at all visits (p<0.05). Thalamic glutamate levels did not differ between groups, but higher levels were related to more pronounced positive symptoms at all visits (p=0.02). The relation between thalamic glutamate levels and negative symptoms was altered over time (negative symptoms*time: p=0.003) due to a significant positive association after two years (p=0.04) but not at other visits. For other metabolites, thalamic NAA were lower in FEP (p=0.04) and total creatine was increased after 6 weeks treatment (p=0.01), whereas dACC glx levels were lower after two years (p=0.02).

The results suggest that greater positive symptom severity is related to higher thalamic glutamate levels and cognitive deficits to lower dACC glutamate levels during the first two years of illness. Furthermore, higher thalamic glutamate levels after two years are associated with more severe negative symptomatology. Findings imply that glutamatergic compounds decreasing thalamic and increasing dACC glutamate levels may be beneficial in FEP over the first two years of illness.

## Introduction

Glutamatergic dysfunction in schizophrenia and psychotic disorders are gradually being acknowledged (1–6), but clinical trials of glutamatergic compounds have overall been negative (7). However, glutamatergic dysfunction and the relationship with clinical symptoms might change over time, and, thus, glutamatergic compounds may mainly be beneficial at specific illness stages.

Schizophrenia and psychotic disorders typically have a clinical onset around the age of twenty (8). This overlaps with the period of prefrontal cortex maturation that is dependent on glutamatergic inputs (9, 10), suggesting glutamatergic dysfunction as a contributing cause of schizophrenia (11). In support, an increasing number of in vivo studies using proton magnetic resonance imaging (1H-MRS) has reported abnormalities in brain levels of glutamate in schizophrenia, and, recently, meta-analyses have been published (1–3, 12). Findings indicate that glutamate levels in anterior cingulate cortex (ACC) and nearby regions are decreased (1, 2, 12) whereas glutamatergic metabolites in the subcortical areas thalamus and basal ganglia are increased when comparing patients in a broad age-range with healthy controls (HC) (2, 3). Studies of antipsychotic-naïve or minimally-treated first-episode patients with psychosis (FEP), on the other hand, suggest a phase-specific pattern in the early course of illness with increased brain levels of glutamate in either thalamus or ACC in patients having a subsequent poor treatment response (4–6, 13). It is therefore likely that glutamate levels change over the course of illness, but this has only been sparsely investigated. A recent four-year follow-up study that investigated FEP within the first two years of illness onset reported a decrease of glutamate levels over time in ACC but not thalamus in both patients and HC (14). Antipsychotic treatment also appears to decrease glutamate levels in the subcortical regions thalamus and striatum, but not hippocampus, in initially antipsychotic-naïve patients receiving first-line treatment (4, 6, 15). In contrast, glutamate levels in ACC seem unaffected after 6 weeks and 6 months treatment (4, 15, 16). Thus, there is a need for longitudinal studies of initially antipsychotic-naïve patients followed-up after longer-term treatment to gain insight into changes in glutamate levels over time as well as the impact of antipsychotic treatment.

Theories of glutamatergic dysfunction have received particular attention by capturing all the three major symptom dimensions of schizophrenia that is positive and negative symptoms and cognitive deficits (17–19). The theory is mainly based on pharmacological challenge studies, genetic- and post mortem studies (19) and needs further validation in patient groups. A recent meta-analysis of 1H-MRS studies reported greater overall symptom severity in patients with lower frontal and higher basal ganglia glutamate levels, but direct associations with positive and negative symptoms were not addressed (2). For positive symptoms, individual studies tend to find higher symptomatology in patients with higher levels of glutamatergic metabolites in FEP (6, 20–23) as well as in the chronic patient populations (24, 25). For negative symptoms, findings are less consistent (5, 6, 13, 21, 26) but greater negative symptom severity in FEP may be associated with higher glutamate levels (5, 13), whereas the reverse pattern has been observed in the chronic phase (27). For cognitive deficits, pharmacological challenge studies and preclinical models imply that glutamatergic dysfunction especially affects the domains attention, spatial working memory (SWM), and executive function (28–32). In support, we previously reported that lower glutamate levels in dACC were associated with impaired performance in tests of attention and spatial working memory but not intelligence in antipsychotic-naïve FEP (33). Other studies have found that both too high (25, 34–36) and too low glutamate levels (37–41) may impair different cognitive domains, but the involvement of glutamate levels in different brain regions on specific cognitive domains as well as the impact of antipsychotics and illness trajectory remains inadequately understood (42).

Last, a range of factors may affect cerebral glutamate levels including age and sex (2, 12, 43), and recent research also points toward the importance of adjustment for measurement quality parameters (1). Furthermore, abnormalities in other metabolites such as reduced frontal myo-inositol (44) and thalamic N-acetylaspartate (NAA) (45) have also been reported in patients with schizophrenia, but the trajectories of these metabolites over two years in initially antipsychotic-naïve patients have not been published.

To address these unknowns, we assessed glutamate levels in dACC and left thalamus in initially antipsychotic-naïve patients with FEP and HC and followed the participants up over two years to relate brain levels of glutamate with positive and negative symptoms and the cognitive domains attention and SWM.

Based on meta-analyses and our previous studies (1–4, 33), we tested the primary hypothesis that glutamate levels in dACC would be reduced in FEP whereas thalamic glutamate levels initially would be higher but reduced after two years when compared with HC. Our second hypotheses explored if lower glutamate levels in dACC was related to a higher degree of cognitive deficits, and if higher glutamate levels in left thalamus were associated with a higher symptom burden of positive symptoms, and if the relation with negative symptoms was altered over the two years. Last, we explored the effect of sex, age at study inclusion, smoking, and dose of antipsychotic compound on glutamate levels.

## Patients and Methods

### Participants and study design

We included 65 antipsychotic-naïve patients with FEP and 57 HC in the Pan European Collaboration on Antipsychotic Naïve Schizophrenia II (PECANSII) study (4, 46) described in detail in the supplementary information. Participants were invited for four visits (baseline, six weeks, six months, and two years) all including a Magnetic Resonance Imaging (MRI) scan, and clinical as well as cognitive assessments as shown in Figure 1.

**Figure 1.**
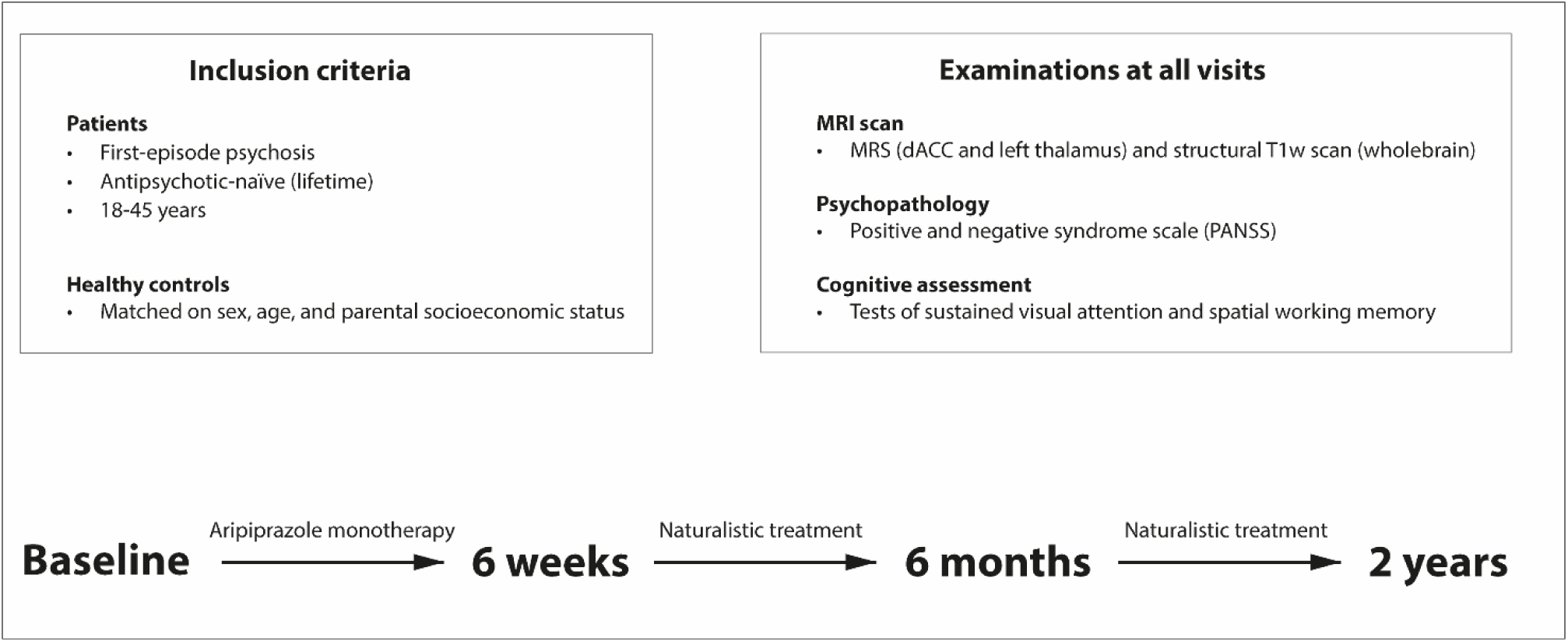
shows the study design for the present study-population. Antipsychotic-naïve first-episode psychosis patients underwent a baseline assessment and were thereafter treated with aripiprazole (tool compounds) as monotherapy until first re-assessment after six weeks. Thereafter, patients could switch to another antipsychotic compound in case of insufficient effect or severe adverse effects before undergoing the third and fourth re-assessments after six months and two years. Aripiprazole serum levels were estimated after six weeks only. HC underwent similar assessments but were not treated with antipsychotics. Participants that dropped out before the six weeks visit were re-invited for the six months and two years visit.

Glutamate levels at baseline, after six weeks and six months have been published previously in overlapping samples (4, 33).

The study was approved by the Committee on Biomedical Research Ethics (H-3-2013-149) and all participants provided written informed consent prior to enrollment in the study. Patients were recruited from in- and outpatient units in the Capital Region of Denmark and the diagnosis was assessed with Schedules for Clinical Assessment in Neuropsychiatry (47) according to the ICD-10 criteria for schizophrenia (DF20.x), schizoaffective disorder (DF25.x) or non-organic psychosis (DF22.x, DF28, or DF29). Antipsychotic-naïve and central stimulant-naïve status (lifetime) was reported by the patients and validated by medical records. HC were recruited via online advertisement (www.forsøgsperson.dk). Exclusion criteria are described in the Supplementary Methods.

### Clinical and cognitive assessment

The Positive and Negative Syndrome Scale (PANSS) (48) and Global Assessment of Functioning scale (49) (social and occupational functioning score (GAF-F)) were used to assess psychopathology and level of function, respectively. Cognitive function was assessed using selected tasks from the Cambridge Neuropsychological Test Automated Battery (50, 51) that was A’ from the rapid visual information processing (RVP) as a measure of sustained visual attention and the strategy score from the SWM task.

### Magnetic Spectroscopy Acquisition, quantification, and quality control

MRI was acquired on a 3.0T Philips scanner (Achieva, Philips Healthcare, Eindhoven, The Netherlands) using a 32-channel head coil (Invivo, Orlando, Florida, USA). First, a structural T1-weighted scan covering the whole brain was acquired for correct anatomical placement of the spectroscopic voxels and tissue segmentation (TR: 10ms; TE: 4.6ms; flip angle: 8°; voxel size: 0.79*0.79*0.80mm^3^). Second, two point-resolved spectroscopy (PRESS) sequences were acquired (TR 3000ms, TE 30ms, 128 averages with MOIST water-suppression, 7 min per scan) together with an inbuild unsuppressed water reference scan in a 2.0×2.0×2.0cm^3^ voxel in dACC (Brodmann area 24 and 32) and a 2.0×1.5×2.0cm^3^ voxel in left thalamus (Supplementary Figure S1 A and C). The magnetic resonance spectroscopy (MRS) spectra were fitted in the spectral range 0.2-4.0ppm using LCModel version 6.3-1L (52) that estimated levels of glutamate, glutamine, glutamate+glutamine (glx), N-acetylaspartate (NAA), total creatine (PCr+Cr), choline, and myo-inositol.

Illustrative glutamate spectra are provided in Supplementary Figure S1 B and D for dACC and left thalamus, respectively. Metabolite levels were quantified in Institutional Units (IU) by adjusting the water-scaled metabolite values for partial-volume cerebrospinal fluid, and gray and white matter using the formula previously described (4, 33) and specified in the Supplementary Information together with the quality control procedure, minimum reporting standards for MRS (53) in Supplementary Table S1, and MRS quality measures in Supplementary Table S2 and S3.

The MRS data in dACC and left thalamus at each visit after quality control are summarized in Figure 2.

**Figure 2.**
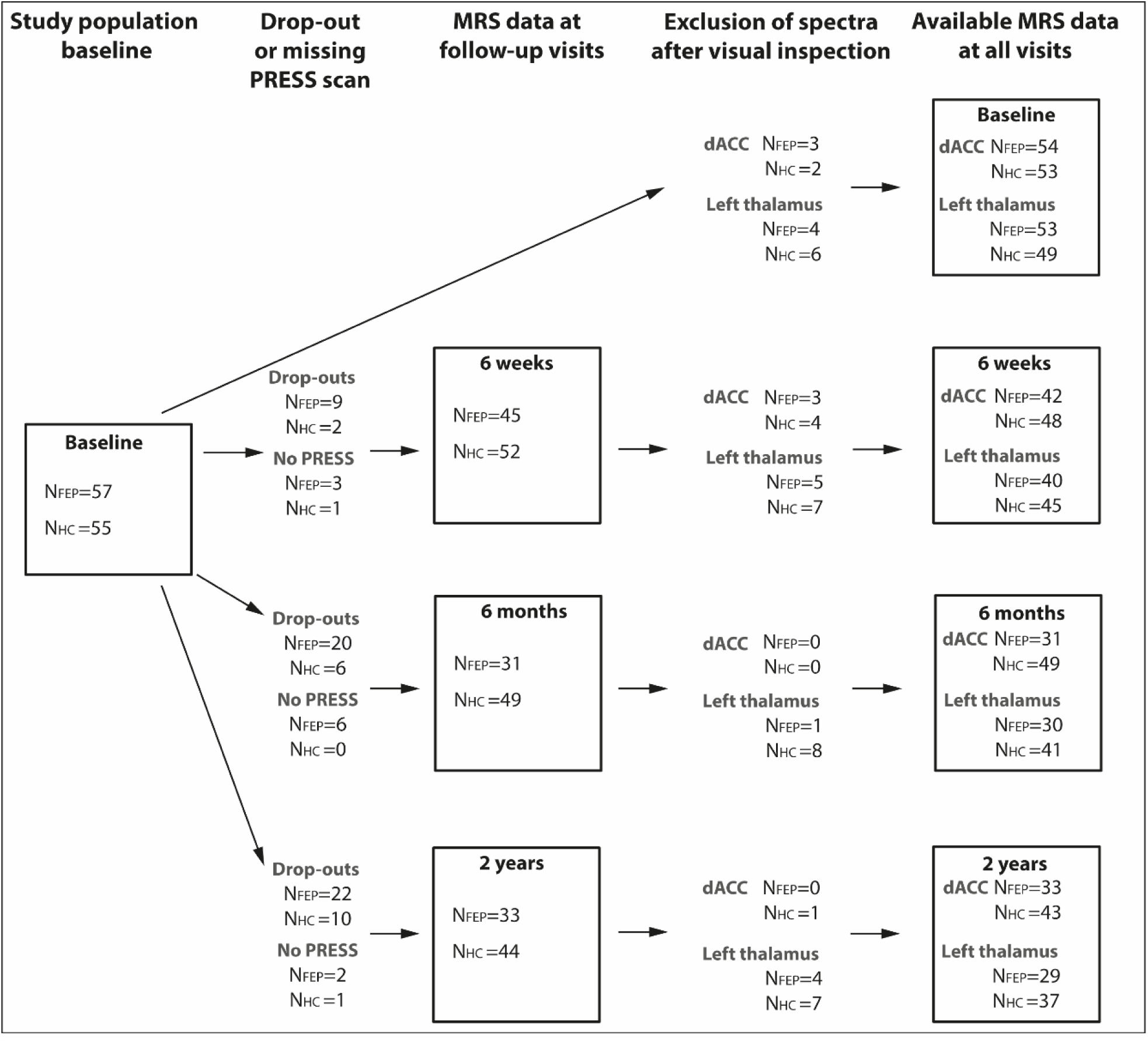
shows the study population at baseline and the number of participants that dropped out or did not undergo a PRESS scan at the three follow-up visits. Exclusion of magnetic resonance imaging spectra and available MRS data for dorsal ACC and left thalamus, respectively, are also shown. Abbreviations: FEP: First-episode psychosis; HC: Healthy control; PRESS: Point Resolved Spectroscopy; MRS: Magnetic resonance spectroscopy; dACC: dorsal Anterior Cingulate Cortex.

### Statistics

Demographic and clinical characteristics were compared between groups using t-tests, *X*^2^, or linear mixed models. SWM strategy scores were log-transformed to conform to normality and multiplied by -1 before statistical analyses so that a higher score indicated better performance.

The trajectory of glutamate and glx over the two years was analyzed using a linear mixed model with group (FEP versus HCs) and time (0, 6, 26, and 96 weeks) as independent variables with adjustment for the nuisance variables age, sex, and smoking status, as well as the scanning quality parameters Cramér-Rao lower bound (CRLB) and Full width at half maximum (FWHM) that differed significantly between FEP and HC (Supplementary Table S2). A significant group*time interaction indicated a different trajectory between groups and was followed-up by post hoc tests at the individual visits. Insignificant interactions were removed, and persistent differences between FEP and HC (main effect of group) or changes over time in both groups (main effect of time) were then evaluated with the p-level set to p<0.025 to adjust for two comparisons (dACC and left thalamus). Explorative main effects of sex, age of participants at study inclusion, and number of cigarettes per day are reported as well.

Similar mixed models were used to explore if the relationship was changed over the two years between glutamate levels and the positive as well as negative score from PANSS, and with antipsychotic dose. For cognitive performance, a similar mixed model also including a group*time*cognitive score interaction first evaluated if there was a significant different trajectory of the association between glutamate levels and cognitive performance in FEP compared with HC. This analysis was followed up by separate mixed models for FEP and HC, respectively.

A general linear model was used to investigate associations between glutamate levels and aripiprazole serum levels after six weeks.

Glx levels are reported as explorative analyses.

The statistical analyses were performed in SAS version 8.3 (SAS institute, Cary, NC, USA).

## Results

For the present study population, eight FEP were excluded due to a missing baseline MRI (due to scanner closedown) and two HC due to brain pathology and later treatment with an antipsychotic compound, respectively, leaving a total of 57 FEP (22.6 ± 5.0 years, 58% females) and 55 HC (22.2 ± 4.3 years) at baseline. After six weeks, 48 FEP and 53 HC were followed-up, 37 FEP and 49 HC after six months, and 35 FEP and 45 HC after two years as shown in Figure 2.

The clinical characteristics and demographics of participants are provided in Table 1. The ethnicity of all HC and 53 out of 57 FEP was white. As expected, FEP and HC differed significantly in years of education (FEP: 12.0+2.1years; HC: 14.2+2.5years; p<0.001), smoking status, cannabis use at the six months visit, and in cognitive performance in sustained visual attention (Table 1). In FEP, the PANSS total score and all sub-scores as well as CGI score decreased significantly after initiation of antipsychotic treatment, and the level of function increased. Chlorpromazine equivalent (CPZE) dose increased at trend-level at the six months and two years visit compared with the six weeks visit (Table 1). The median of duration of psychosis for FEP was 27 weeks (25-75^th^ percentile: 15-117 weeks), mean dose of aripiprazole after six weeks was 10.2±4.8mg, and mean serum level 123.0±66.0µg/L.

**Table 1:**
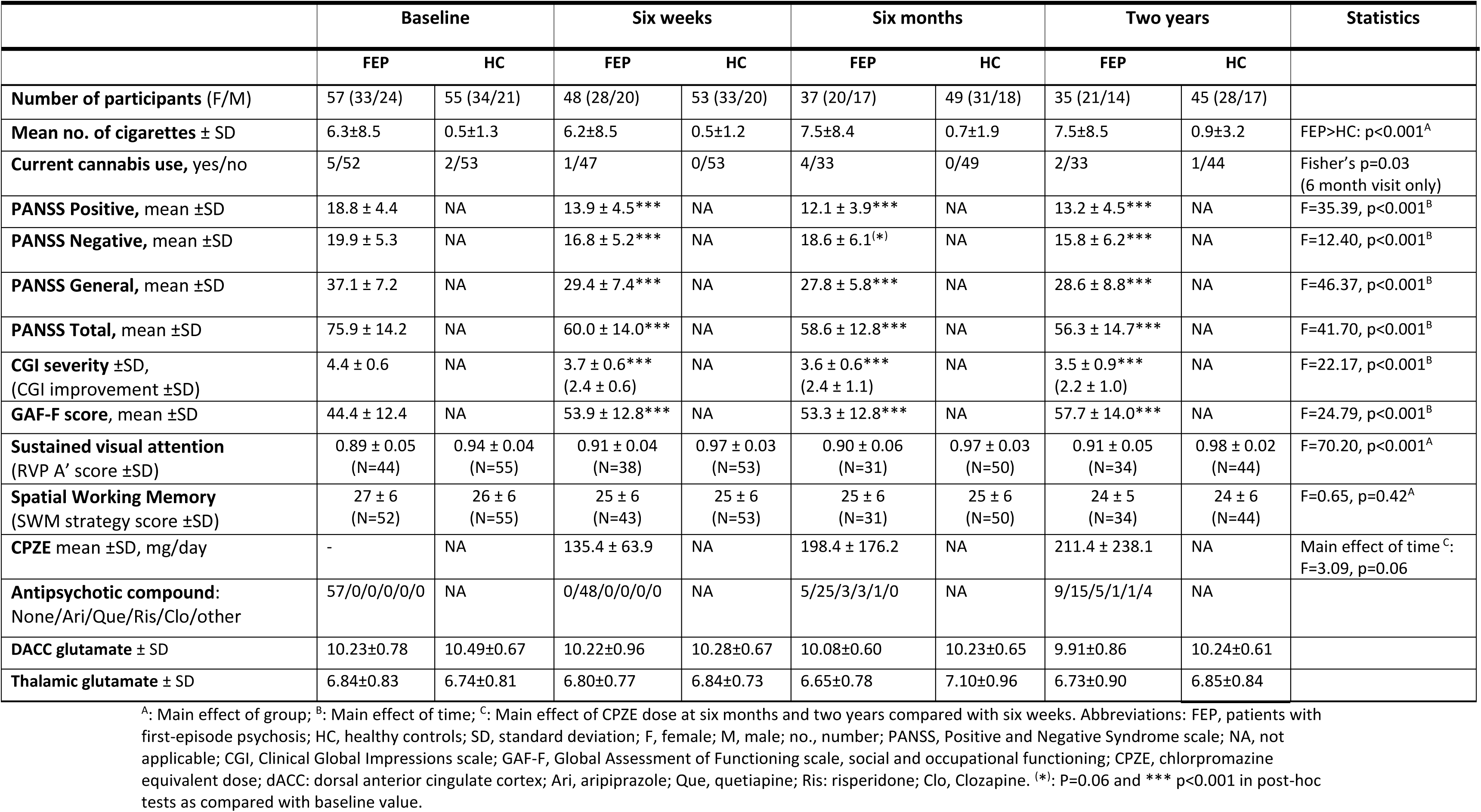
Demographics and Clinical Characteristics of Participants and mean glutamate levels

### Glutamate levels over two years in patients compared with healthy controls

The trajectory of glutamate levels in dACC and left thalamus over the two years in FEP compared with HC is illustrated in Figure 3, statistics are reported in Table 2, and findings are briefly summarized below. The mean levels of glutamate are provided in Table 1 and glutamate and glx levels as well as and gray- and white-matter fraction in the dACC and left thalamus spectroscopic voxels in supplementary Table S4.

**Figure 3.**
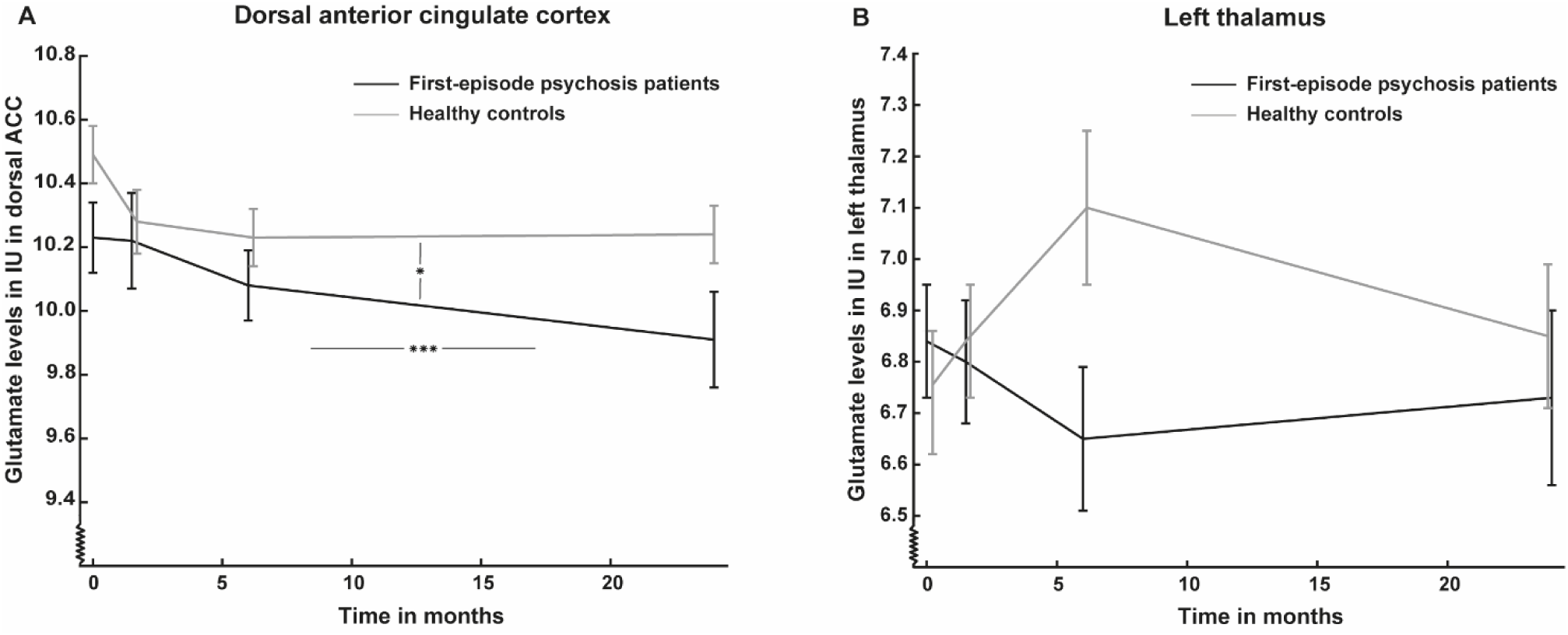
shows mean glutamate levels and standard errors in (A) dorsal anterior cingulate cortex (dACC) and (B) left thalamus over two years in the initially antipsychotic-naïve first-episode psychosis patients (FEP) (black lines) compared with healthy controls (HC) (gray lines). Participants were assessed at baseline (0 months), after six weeks (1.5 months), six months, and two years (24 months). A: Glutamate levels in dACC were lower in FEP at all visits and decreased over two years in both FEP and HC. B: Glutamate levels did not differ between FEP and HC over the two years and were not changed over time. Vertical bar represents main effect of time, and horizontal bar represents main effect of group. *: p<0.05 and ***: p<0.001.

**Table 2:**
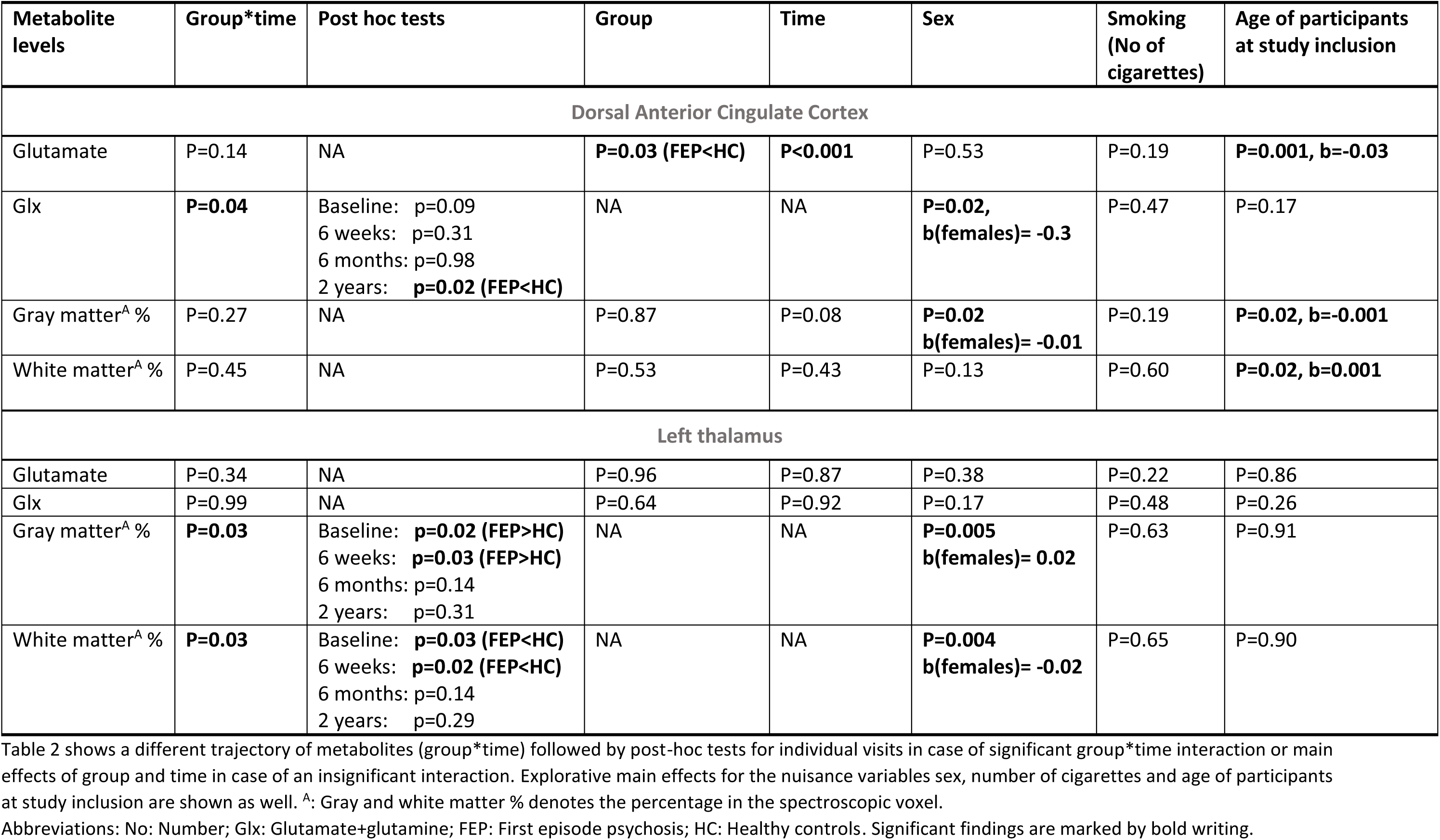
Differences in the trajectory or main effects of glutamatergic metabolites over two years in first-episode patients and healthy controls

#### Dorsal ACC

The trajectory of glutamate levels did not differ between FEP and HC over the two years, but glutamate levels were significantly lower in FEP compared with HCs at all visits and decreased significantly over time in both groups (Table 2). This may suggest that dACC glutamate levels are lower in FEP and that the age-related decrease is not accelerated in the patients over the first two years of the illness.

The trajectory of glx levels was significantly different in FEP compared with HC due to lower glx levels in FEP after two years (Supplementary Figure S2A), which may suggest an accelerated decrease in FEP after two years illness.

#### Left thalamus

The trajectory of glutamate and glx levels did not differ significantly over the two years, and the main effects of group and time were insignificant.

### Levels of other metabolites over two years in patients compared with healthy controls

The trajectories of other metabolite levels than glutamate, which differed significantly over the two years in FEP compared with HC or were changed over time in both groups, are illustrated in Supplementary Figure S2 and S3, mean levels are reported in Supplementary Table S5, and statistics are summarized in Supplementary Table S6.

#### Dorsal ACC

NAA and tCr levels significantly decreased in both groups over the two years (Supplementary Figure S2B and C, and Supplementary Table S6). For other metabolite levels than NAA and tCr, the trajectory did not differ between FEP and HC over the two years, and main effects of group and time were insignificant.

#### Left thalamus

For NAA, there was a significant main effect of group due to lower levels in FEP over the two years (Supplementary Table S6 and Figure S3A). For myo-inositol and tCr, there was significant different trajectories in FEP compared with HC (Supplementary Table S6). For myo-inositol, there was significantly higher levels in FEP after two years and for tCr, there were significantly higher levels in FEP after six weeks treatment and a non-significant trend for higher levels after six months (Supplementary Figure S3B and C).

#### The effect of sex, smoking, and age of participants

There were significant main effects of sex for glx levels in dACC due to lower values in females, and significant main effects of age of participants at study inclusion for dACC glutamate levels due to a decrease with increasing age. Smoking did not affect glutamate and glx levels significantly.

For other metabolites, choline levels in dACC and left thalamus were lower in females, and there were borderline significant higher levels of dACC tCr in smokers. For age at study inclusion, there was higher levels of dACC myo-inositol and choline as well as thalamic myo-inositol and tCr with increased age.

### Glutamate levels over two years in patients and healthy controls analyzed separately

#### Dorsal ACC

When analyzing FEP only, there was a significant main effect of time for both glutamate (p<0.001) and glx (p=0.01) due to a decrease over the two years, whereas the main effect of time in the HC group analyzed separately was insignificant (p=0.10-0.18).

#### Left thalamus

The main effect of time was insignificant for FEP analyzed separately (p=0.86) and also for HC analyzed separately (p=0.45).

Other metabolites over two years in patients and healthy controls analyzed separately are reported in the supplementary results.

### The association between glutamatergic metabolites and psychopathology over two years

#### Dorsal ACC

There were no significant associations between glutamate or glx levels and the PANSS positive and negative scores over the two years (Table 3). Moreover, the association between glutamatergic metabolites and clinical symptoms did not significantly change over the two years (PANSS score*time insignificant) (Table 3).

**Table 3:** The association between glutamatergic metabolite levels and clinical outcome in first-episode patients over two years

|  | Clinical variable*time | Post hoc tests | Main effect clinical variable |
| --- | --- | --- | --- |
| **Dorsal ACC glutamate levels** |  |  |  |
| PANSS-positive | P=0.71 | NA | P=0.62 |
| PANSS-negative | P=0.34 | NA | P=0.60 |
| Sustained visual attention | P=0.11 | NA | **P<0.05, b=2.3** |
| Spatial working memory | P=0.67 | NA | P=0.55 |
| **Dorsal ACC glx levels** |  |  |  |
| PANSS-positive | P=0.59 | NA | P=0.50 |
| PANSS-negative | P=0.59 | NA | P=0.71 |
| Sustained visual attention | **P=0.03** | Baseline: p=0.07<br>6 weeks: p=0.61<br>6 months: p=0.27<br>2 years: p=0.27 | NA |
| Spatial working memory | P=0.30 | NA | P=0.27 |
| **Left thalamus glutamate levels** |  |  |  |
| PANSS-positive | P=0.15 | NA | **P=0.02, b=0.04** |
| PANSS-negative | **P=0.003** | Baseline: **p=0.06, b=-0.04**<br>6 weeks: p=0.17<br>6 months: p=0.39<br>2 years: **p=0.04, b=0.04** | NA |
| Sustained visual attention | P=0.25 | NA | P=0.86 |
| Spatial working memory | P=0.25 | NA | P=0.60 |
| **Left thalamus glx levels** |  |  |  |
| PANSS-positive | P=0.63 | NA | **P=0.04, b=0.04** |
| PANSS-negative | P=0.23 | NA | P=0.16 |
| Sustained visual attention | P=0.78 | NA | P=0.79 |
| Spatial working memory | **P=0.02** | Baseline: p=0.65<br>6 weeks: p=0.22<br>6 months: p=0.18<br>2 years: p=0.07, b=4.9<sup>A</sup> | NA |

Table 3 shows associations between glutamatergic metabolite levels and clinical symptoms. A significant clinical variable \*time interaction indicates a change in the association over two years and is followed up by post-hoc tests for individual visits. The main effect of the clinical variable is reported in case of an insignificant interaction.
Abbreviations: PANSS: Positive and Negative Syndrome scale; ACC: Anterior cingulate cortex; glx:
Glutamate+glutamine; NA: not applicable. <sup>A</sup>: Spatial working memory strategy score was log-transformed to obtain normality and multiplied by -1 prior to statistical analyses so that higher score indicated improved performance.

#### Left thalamus

There was a significant positive association between the PANSS positive score and glutamate as well as glx levels over the two years, but not a different association between glutamatergic metabolites and psychotic symptoms over time (PANSS positive score*time insignificant) (Table 3). For the PANSS negative score, the association between thalamic glutamate levels and negative symptoms was changed over the two years (PANSS negative*time: p=0.003), which was due to a trend-level negative association at baseline but a significant positive association after two years (Table 3). A similar pattern was not found for glx (Table 3).

### The association between glutamatergic metabolites and cognitive domains over two years

#### Dorsal ACC

##### Attention

There was a significant different trajectory of the association between glutamate levels and RVP A’ score over the two years in FEP and HCs (group*time*RVP A’ score: p=0.026) due to a positive association between glutamate levels and RVP A’ score over the two years in the FEP group (Table 3) but not in the HCs (main effect: p=0.41). A similar trend was seen for glx levels and RVP A’ score.

##### Spatial working memory

The association between glutamate levels and spatial working memory strategy score did not differ over the two years in FEP and HC (group*time*SWM strategy score: p=0.42) and there was no significant main effect of SWM strategy in FEP (Table 3) or HC (p=0.73).

#### Left thalamus

The associations between glutamate levels and cognitive scores did not differ over the two years in FEP and HC (group*time*cognitive score: p=0.25-0.99), but in FEP there was a different association between glx levels and SWM strategy due to a trend for higher glx levels being associated with better performance after two years but not at any other visit (Table 3). In the HC, there was a different association between glx levels and RVP A’ score over the two years (time*RVP A’ score: p=0.04) due to higher glx levels being associated with better cognitive performance after two years (b=17.2, p=0.04) but not at any other visit (p=0.10-0.79).

##### The association between glutamatergic metabolites and dose of antipsychotics at the follow-up visits Dorsal ACC

There was a significant different association between CPZE dose at the follow-up visits and glutamate levels (CPZE*time: p=0.03) due to a trend for a positive association between glutamate levels and CPZE at six weeks (b=0.003, p=0.08) but not after six months (p=0.11) or two years (p=0.24). Similarly, a borderline significant different association was found for glx levels (CPZE*time: p=0.05) due to a borderline significant positive association after six months (b=0.001, p=0.05) but not after six weeks (p=0.12) or two years (p=0.24). The serum level of aripiprazole after six weeks was not associated with glutamate (p=0.12) or glx levels (p=0.82).

##### Left thalamus

CPZE dose and glutamate or glx levels levels were not associated at any follow-up visits (main effect: p=0.33-0.74), but serum aripiprazole after six weeks was significantly positively associated with glutamate levels (b=0.005, p=0.02) and borderline significantly with glx levels (b=0.007, p=0.05).

### The association between glutamatergic metabolites in dACC and left thalamus over two years

Exploratory analyses revealed that the association between glutamate levels in dACC and left thalamus changed over the two years, which seemed to be due to a positive association between glutamate levels in dACC and left thalamus in the combined group of FEP and HC after six weeks but a non-significant trend for a negative association after two years as described in the Supplementary Information and Figure S4.

### Drop-out analyses

There were no significant baseline differences between patients that participated and dropped-out from the two-year visit when comparing glutamate levels and PANSS positive or negative sub-score (Supplementary Table S7).

## Discussion

To our knowledge this is the first study to report on glutamate levels over two years in initially antipsychotic-naïve FEP and relate metabolite levels to both positive and negative symptoms as well as cognitive deficits. Main findings are that dACC glutamate levels were significantly lower in FEP and related to attentional deficits, and that thalamic glutamate levels were related to positive and negative symptoms.

### Glutamate levels over two years

In dACC, we found significant lower glutamate levels in FEP compared with HC at all visits, which extends findings from recent meta-analyses (1–3) and longitudinal studies of medicated or minimally treated FEP (14, 16) by showing that decreased cortical levels are present from illness onset. We did not find an accelerated decrease of dACC glutamate levels as suggested by preclinical studies (11) and the first meta-analysis of glutamate levels in schizophrenia (54) but not confirmed by recent meta-analyses (1, 12, 55). It is possible, though, that an accelerated decrease of glutamatergic metabolites appears later in the illness phase. First, we observed a significant decrease of glutamate levels over the two years in FEP but not in HC, when the groups were analyzed separately, and, secondly, we observed decreased glx levels after two years in FEP but not HC. Thus, studies with longer follow-up times are warranted.

In left thalamus, we observed no group differences in glutamate levels at baseline, which contrasts our previous finding of increased glutamate levels in the antipsychotic-naïve state being reduced after first-line treatment (4, 33). Several reasons might explain these discrepancies. First, we used glutamate calculated as IU in the current study as opposed to creatine-scaled values in a previous study (4). Second, increased thalamic glutamate levels may characterize the most severely ill patients as we previously reported increased thalamic glutamate levels in IU in antipsychotic-naïve patients with a schizophrenia diagnosis, but not in a combined sample of patients with schizophrenia and psychotic disorders (33), and it is likely that the most severely ill patients dropped-out from the two years visit. In line with this, other studies have also reported that increased glutamate levels mainly are found in patients with a higher symptom burden (13, 56, 57). Last, the lack of increased glutamate levels at baseline in thalamus may be due to a power issue as signal-to-noise ratio (SNR) was lower and CRLB values higher in left thalamus compared with dACC.

Interestingly, the association between glutamate levels in dACC and left thalamus appeared to be similar in FEP and HC after six weeks but not at baseline. This may reflect that first-line treatment may restore deficits in cortico-striato-thalamo-cortical networks suggested to lead to cognitive deficits and symptoms of schizophrenia (58–60).

### Glutamate levels and clinical symptoms over the two years

Thalamic glutamate levels were related to positive and negative symptoms, whereas glutamate levels in dACC were associated with cognition in a test of sustained attention. Of these findings, the relationship with negative symptoms was dynamic, whereas associations with positive symptoms and cognitive deficits were stable over the two years.

The association between higher thalamic glutamate levels and PANSS positive score over two years extends our previous finding of a relationship between thalamic glutamate levels in antipsychotic-naïve patients with subsequent non-response to first-line antipsychotic treatment in an overlapping cohort (4) and is in accordance with another study finding increases in thalamic glx in minimally treated FEP not responding to first-line treatment (22). Moreover, we also observed an association between higher serum-levels of antipsychotics and higher thalamic glutamate levels after six weeks. Thus, higher thalamic glutamate levels between patients may be a marker for a more severe illness trajectory over the first two years of illness.

The dynamic relationship between thalamic glutamate levels and negative symptoms may indicate that at baseline higher glutamate levels were beneficial for negative symptoms severity, whereas the reverse pattern was found after two years. In support, cross-sectional studies imply that a similar dynamic relationship over time may exist for ACC glutamate levels and negative symptoms due to a positive association in FEP (5, 13) but a negative association in a chronic patient population (27). Future studies should therefore aim for reproduction of these findings in larger samples, preferentially in multi-center studies. This is clinically relevant, since emerging evidence suggests an effect of glutamate modulating agents on negative symptoms (61). Illness phase specific prescription guidelines of glutamate modulating compounds may be needed, if the relationship between glutamate levels and negative symptoms is altered over the course of illness.

Lower glutamate levels in dACC in FEP were associated with a higher degree of cognitive deficits in a test of attention over the two years, which extends our previous finding of a similar association in antipsychotic-naïve FEP (33). A recent cross-sectional study of unmedicated and medicated patients found that treatment may alter the association between brain glutamate levels and cognitive function (62), but since we found a stable association over two years, our findings suggest that this presumed alteration appears later in the illness phase.

The mechanism behind the correlation between resting levels of glutamate in dACC and cognition is incompletely understood, but recent functional MRS studies have shown perturbations in a transient up-regulation of ACC glutamate levels in patients compared with HC performing a cognitive task in a scanner (63, 64). It is possible that the lower resting dACC glutamate levels we found in FEP makes the patients more vulnerable for such a deficit in glutamatergic upregulation.

We did not, however, find associations between dACC glutamate levels and positive or negative symptoms. However, although findings did not support direct relationships between dACC glutamate levels and specific symptom clusters, other studies have reported an association between higher ACC glutamate levels and poor response to treatment (5, 6, 13) although not consistently observed (4, 65, 66). The differences may be due to voxel placement. We placed our voxel in dorsal ACC, but higher levels of glutamate are in general found in more ventral parts of ACC in both FEP (5, 6, 13) and HC (67) compared with more dorsal regions (4).

Interestingly, our findings indicate a reverse relationship between glutamate levels in a cortical and a subcortical area and symptomatology in FEP. More specifically, lower dACC levels of glutamate were related to a higher degree of cognitive deficits, whereas higher thalamic glutamate levels were associated with more severe positive and negative symptom severity, except for negative symptoms at baseline. This is in accordance with a recent meta-analysis reporting a relation between higher PANSS total score and lower frontal as well as higher striatal glutamate levels (2). These combined findings may suggest that lower cortical and higher subcortical glutamate levels are part of the schizophrenia pathophysiology with subcortical levels being related to mainly psychotic symptoms and cortical levels with mainly cognitive performance. This pattern mimics the dopamine hypothesis of schizophrenia postulating increased subcortical dopaminergic activity being related to psychosis and reduced prefrontal dopaminergic activity underlying cognitive deficits (68, 69). It may imply that glutamatergic therapies with an effect similar to partial dopamine agonism (70) with increases in areas with too low activity but decreases in areas with too high could be favorable in FEP.

Intriguingly, the relationship between glutamate levels in dACC and left thalamus appeared to change over the two years (Supplementary Figure S4), and this may impact the relationship between clinical symptoms and glutamate levels at a later illness stage.

### The effect of sex and age at study inclusion

Sex differences in MRS studies are sparsely studied, but existing evidence suggests lower prefrontal glutamate levels in females in HC (43). Furthermore, a recent study reported lower glutamate in dACC in genetically high risk (GHR) of schizophrenia females compared with GHR males (71). In support, we found lower levels of glx in dACC in females. This may have implications for interpretation of findings in MRS studies. For example, in the current study sample, we had an overweight of 58% females compared to a proportion of 70-80% males in the majority of other MRS studies of antipsychotic-naïve or minimally-treated FEP (5, 6, 65). Thus, differences in sex distribution between studies may partly explain why some find increased levels of glutamatergic metabolites in prefrontal regions in non-responders (5, 6) whereas others do not (4).

Higher age at study inclusion was significantly associated with lower glutamate levels in dACC as well. The decrease of glutamate levels in dACC with increasing age is in line with some meta-analyses (12, 54) although not all (1).

### The trajectory of other metabolites than glutamate

Thalamic levels of NAA were significantly lower over the two years in FEP and unaffected by longer-term treatment, which is in line with a meta-analysis (45) and studies reporting no effect of short-term treatment on thalamic NAA levels (22, 73).

Interestingly, thalamic tCr increased after first-line treatment at six weeks and at trend level after six months but normalized after two years. This pattern mimics our previous findings of increased striatal perfusion after six weeks and six months but not after two years in an overlapping cohort (74, 75). These findings suggest a transient increase in subcortical metabolism (76, 77) after first-line treatment that seems to normalize after two years.

Last, thalamic myo-inositol increased over the two years in FEP, which should be investigated further given that higher myo-inositol levels are found in treatment resistant schizophrenia (78).

A major strength of the current study is recruitment of a large antipsychotic-naïve FEP group that was followed up over a long period, whereas the greatest limitation was a drop-out rate after two years on 39%. Moreover, glutamate levels were quantified on a 3T scanner, where glutamate and glutamine are not as reliably separated as compared with higher field strengths such as 4T and 7T, and measures of glutamate levels in the present study are therefore contaminated to some degree by the glutamine (79).

Last, we had an overweight of females, which should be taken into account when comparisons with other studies are made.

## Conclusions

The present study suggests that dACC levels of glutamate are lower in initially antipsychotic-naïve FEP over the first two years of illness and do not decline at an accelerated rate in patients in this period. Moreover, greater symptom severity of positive symptoms and cognitive deficits is related to higher thalamic and lower dACC glutamate levels over the first two years in a stable manner, whereas the relationship with negative symptoms seems dynamic. The findings indicate a potential for glutamate compounds that decrease thalamic and increase dACC glutamate levels in FEP in the first two years of illness.

## Data Availability

All data produced in the present study may be available upon reasonable request to the authors through a formal collaboration agreement

## Acknowledgements

This study was funded by an independent grant from the Lundbeck Foundation (R155-2013-16337) for the Lundbeck Foundation Centre of Excellence for Clinical Intervention and Neuropsychiatric Schizophrenia Research (CINS) (B Glenthøj); and a Ph.D. grants from the Faculty of Health and Medical Sciences, University of Copenhagen (KB Bojesen) as well as a post doc grant from the research Fund 2022 in the Capital Region of Denmark (KB Bojesen); grants from the Wørzner (B Glenthøj) and Gerhard Linds Foundations (KB Bojesen); and support from the Mental Health Services, Capital Region of Denmark (B Glenthøj).

*Role of the Funders/Sponsors*: The funding sources had no role in the design and conduct of the study; collection, management, analysis, and interpretation of the data; preparation, review, or approval of the manuscript; or decision to submit the manuscript for publication.

