## Supplementary information for "Cerebral glutamate levels over two years in initially antipsychotic-naïve first-episode patients with psychosis are related to clinical symptoms and cognition"

### Supplementary Methods

*Study Design*

Participants are part of the multimodal longitudinal Pan European Collaboration on Antipsychotic Naïve Schizophrenia II (PECANSII) study (1, 2) that investigated initially antipsychotic-naïve patients with psychosis at baseline and followed patients up after 6 weeks of monotherapy with aripiprazole. Patients were further re-invited for re-examination after 6 months and 2 years of naturalistic treatment (clinicians' choice). Healthy controls (HCs) matched on age, sex, and parental socioeconomic status were recruited through online advertisement and underwent similar assessments as patients but did not receive antipsychotics or any other treatment. Participants were examined with magnetic resonance imaging (MRI) including proton magnetic resonance spectroscopy (1H-MRS) as well as structural and functional MRI sequences, neurocognitive testing, electrophysiological examinations, blood sampling, and a subgroup of participants also had a 3,4-dihydroxy-6-[18F] fluor-L-phenylalanine ($^{18}$F-DOPA) positron emission tomography (PET) scan at baseline and after 6 weeks (3).

Glutamate levels were estimated with single voxel MRS in dorsal anterior cingulate cortex (dACC) and left thalamus. The MRS regions-of-interests (ROIs) were chosen since they are connected in a cortico-striato-thalamo-cortical network presumed to underlie psychotic disorder (4) and play a key role in the schizophrenia pathophysiology (5, 6). The left thalamus was chosen as ROI instead of the right thalamus since previous studies mainly had reported on abnormalities in glutamatergic metabolites in left thalamus in the planning phase of the PECANSII study (7).

*Participants*

Exclusion criteria for antipsychotic-naïve patients with first episode psychosis (FEP) only were treatment with an antidepressant within the last 30 days and being involuntarily admitted or treated, and for healthy controls (HC) only it was having a psychiatric or physical disease or a first degree relative with psychiatric illness. Further exclusion criteria for all participants were substance abuse (corresponding to a DF10.x diagnosis), previously head injury with more than five minutes of unconsciousness, pregnancy, and severe physical illness as well as contradictions for a MRI scan (pacemaker and certain metallic implants).

*Magnetic Spectroscopy Acquisition, quantification, and quality control*

*Quantification*: Water-scaled metabolite values were adjusted for partial-volume cerebrospinal fluid and gray as well as white matter fractions to calculated metabolite level in institutional units ($M_{IU}$) in dorsal anterior cingulate cortex (dACC) and left thalamus by using the following formula (8):

$$M_{IU} = M*(WM+GM+1.55*CSF)/(WM+GM)$$

$M_{IU}$: Metabolite level in institutional units
M: In vivo water-scaled values of metabolites from LCModel output
WM: White matter fraction in the spectroscopic voxels
GM: Gray matter fraction in the spectroscopic voxels
CSF: Cerebrospinal fluid content in the spectroscopic voxels (1-WM+GM)

*Minimum reporting standards for in vivo magnetic resonance spectroscopy*

Minimum reporting standards for in vivo magnetic resonance spectroscopy are reported in supplementary Table S1 (9).

**Supplementary Table S1: Minimum reporting standards for in vivo Magnetic Resonance Spectroscopy**

| Site (name) | Center for Neuropsychiatric Schizophrenia Research (CNSR), Mental Health Center Glostrup, Denmark |
|---|---|
| 1. Hardware | |
| a. Field strength [T] | 3T |
| b. Manufacturer | Philips Achieva |
| c. Model (software version if available) | Philips dSTREAM Achieva |
| d. RF coils: nuclei (transmit/receive), number of channels, type, body part | 32-channel head coil |
| e. Additional hardware | None |
| 2. Acquisition | |
| a. Pulse sequence | PRESS |
| b. Volume of interest (VOI) locations | Dorsal anterior cingulate cortex (dACC) (Supplementary Figure S1A)<br>Left thalamus (Supplementary Figure S1C) |
| c. Nominal VOI size [cm3] | dACC: 2.0*2.0*2.0cm$^3$<br>Left thalamus: 2.0*1.5*2.0cm$^3$ |
| d. Repetition time (TR), echo time (TE) [ms] | TR=3000ms, TE=30ms |
| e. Total number of excitations or acquisitions per spectrum | 128 averages |
| f. Additional sequence parameters (spectral width in Hz, number of spectral points, frequency offsets) | 2500 Hz, 1024 samples |
| g. Water suppression method | MOIST |
| h. Shimming method, reference peak, and thresholds for “acceptance of shim” chosen | Second order Pencil Beam auto |
| i. Triggering or motion correction method (respiratory, peripheral, cardiac triggering, incl. device used and delays) | None |

| 3. Data analysis methods and outputs | |
|---|---|
| a. Analysis software | LCModel version 6.3-1L |
| b. Processing steps deviating from quoted reference or product | None |
| c. Output measure (eg absolute concentration, institutional units, ratio), processing steps deviating from quoted reference or product | Metabolite concentrations adjusted for partial-volume cerebrospinal fluid and the fraction of gray and white matter in the voxel.<br>No processing steps deviated from quoted references or product. |
| d. Quantification references and assumptions, fitting model assumptions | LCModel basis set<br>press_te30_3t_gsh_v3 |
| **4. Data quality** | |
| a. Reported variables (SNR, linewidth (with reference peaks)) | SNR and FWHM (Supplementary Table S2) |
| b. Data exclusion criteria | Spectra were excluded after visual inspection according to the 'Criteria for rejecting analyses' in the LCModel & LCMgui User's Manual (http://s-provencher.com/pub/LCModel/manual/manual.pdf), Chapter 3.5.<br>Individual metabolite values were excluded if Cramér-Rao lower bound (CRLB) > 20% |
| c. Quality measures of postprocessing model fitting (eg CRLB, goodness of fit, SD of residual) | CRLB (Supplementary Table S2) |
| d. Sample spectrum | dACC: Voxel location shown in Supplementary Figure S1B<br>Left thalamus: Voxel location shown in Supplementary Figure S1D |

Supplementary Table S1 shows minimum reporting standards for in vivo magnetic resonance spectroscopy. Abbreviations: PRESS: point-resolved spectroscopy; dACC: Dorsal anterior cingulate cortex; Hz: Hertz; VOI: volume of interest; TR: Repetition time; TE: echo time; MOIST: multiple optimizations insensitive suppression train; SNR: Signal-to-noise ratio; CRLB: Cramér-Rao lower bound; FWHM: Full-width half-maximum.

**Supplementary Figure S1**

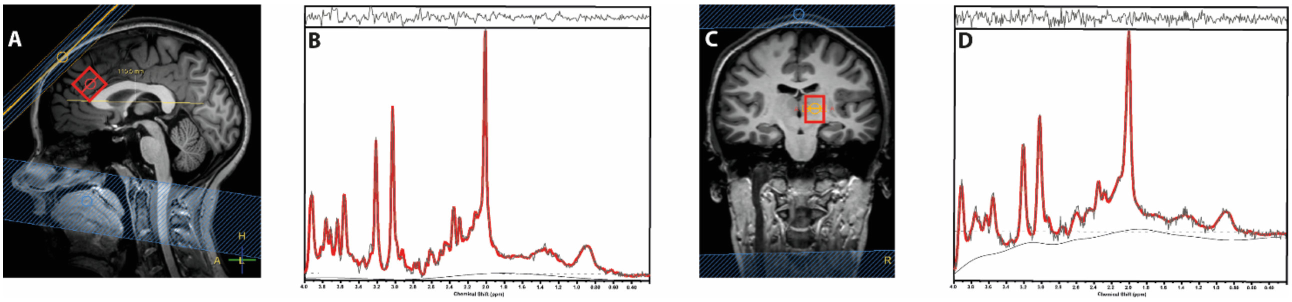


Supplementary Figure S1 shows voxel location in dorsal anterior cingulate cortex (dACC) (A) and left thalamus (C) as well as representative spectra from dACC (B) and left thalamus (D). The top bar of the spectra shows residuals. Abbreviations: ppm: parts per million.

*Magnetic Spectroscopy resonance quality control*

Quality control was done by first evaluating all spectra by using the 'criteria for rejecting analyses' in the LCModel & LCMgui User's Manual (http://s-provencher.com/pub/LCModel/manual/manual.pdf), chapter 3.5. For individual metabolites, the exclusion criterium was a Cramér-Rao lower bound (CRLB) > 20%. In dACC, CRLB $\leq$ 20% for glutamate, glx and all other metabolites than glutamine (gln) at all visits, whereas exclusion due to CRLB > 20% was done for gln (baseline: 9 FEP and 11 HCs; 6 weeks: 7 FEP and 4 HCs; 6 months: 8 FEP and 7 HCs; 2 years: 8 FEP and 7 HCs). In left thalamus, exclusion due to CRLB > 20% were done for one glutamate data in one HC at baseline only, for one myo-inositol data in one HC at the two-years visit, and for gln data in 49 FEP and 42 HC at baseline, why gln data at all visits were excluded from the analyses. For the remaining metabolites, all CRLB $\leq$ 20% at all visits in left thalamus. Full width at half maximum (FWHM), signal-to-noise ratio (SNR), and CRLB-values for glutamate and glx are provided in Supplementary Table S2 reporting significantly higher FWHM and CRLB-values for glutamate in dACC in FEP, but no group differences in left thalamus. CRLB-values for other metabolites are provided in Supplementary Table S3.

**Supplementary Table S2: Quality measures in dorsal Anterior Cingulate Cortex and left thalamus**

| **Dorsal anterior cingulate cortex** | | | | | | | | | |
|---|---|---|---|---|---|---|---|---|---|
| | **Baseline** | | **6 weeks** | | **6 months** | | **2 years** | | **Statistics** |
| | **FEP** | **HC** | **FEP** | **HC** | **FEP** | **HC** | **FEP** | **HC** | **Main effect of group** |
| **N** | n=54 | n=53 | n=42 | n=48 | n=31 | n=49 | n=33 | n=43 | |
| FWHM | 0.030±0.008 | 0.0026±0.006 | 0.029±0.008 | 0.0027±0.005 | 0.031±0.008 | 0.0027±0.006 | 0.029±0.006 | 0.028±0.006 | **P=0.01** |
| SNR | 31.7±3.0 | 32.0±2.9 | 31.0±3.6 | 31.9±2.6 | 31.0±3.3 | 31.8±2.5 | 31.8±2.8 | 31.7±2.2 | P=0.61 |
| CRLB (%) Glu | 5.2±0.6 | 5.1±0.5 | 5.3±1.0 | 5.1±0.3 | 5.3±0.4 | 5.1±0.3 | 5.2±0.5 | 5.0±0.3 | **P=0.03** |
| CRLB (%) Glx | 5.0±0.8 | 4.9±0.6 | 5.1±1.0 | 4.9±0.6 | 5.2±0.6 | 4.9±0.6 | 5.1±0.6 | 4.9±0.5 | P=0.10 |
| **Left thalamus** | | | | | | | | | |
| **N** | n=53 | n=49 | n=40 | n=45 | n=30 | n=41 | N=29 | N=37 | |
| FWHM | 0.047±0.007 | 0.047±0.007 | 0.049±0.012 | 0.046±0.005 | 0.046±0.007 | 0.048±0.006 | 0.048±0.008 | 0.045±0.007 | P=0.61 |
| SNR | 16.2±3.6 | 16.6±3.4 | 16.8±4.7 | 16.5±3.3 | 17.0±3.1 | 16.3±3.4 | 16.5±3.2 | 18.0±4.3 | P=0.78 |
| CRLB (%) Glutamate | 9.8±1.8 | 9.8±1.8, n=48[A] | 10.1±1.6 | 9.8±1.4 | 9.8±1.6 | 9.3±1.2 | 9.9±1.5 | 9.5±1.8 | P=0.41 |
| CRLB (%) Glx | 8.6±1.3 | 8.7±1.6 | 8.9±1.3 | 8.5±1.4 | 8.7±1.8 | 8.4±1.2 | 8.9±1.4 | 8.8±2.1 | P=0.72 |

Abbreviations: ACC: Anterior cingulate cortex; FEP: First episode psychosis; HC: Healthy controls; FWHM: Full-width half-maximum; SNR: Signal-to-noise ratio; CRLB: Cramér-Rao lower bound; Glx: Glutamate+glutamine.

Group differences in quality measures during the two years were tested with linear mixed models. The group*time interaction terms were insignificant for all CRLB values, FWHM and SNR in dACC and left thalamus, and were removed from the statistical models.

**Supplementary Table S3: Quality measures of other metabolites in dorsal Anterior Cingulate Cortex and left thalamus**

| | Baseline | | 6 weeks | | 6 months | | 2 years | | Statistics |
|---|---|---|---|---|---|---|---|---|---|
| | **FEP** | **HC** | **FEP** | **HC** | **FEP** | **HC** | **FEP** | **HC** | **Main effect of group** |
| **Dorsal Anterior Cingulate Cortex** | | | | | | | | | |
| N | n=54 | n=53 | n=42 | n=48 | n=31 | n=49 | n=33 | n=43 | |
| CRLB (%) Gln | 16.5±2.4, n=45[A] | 16.4±2.2, n=42[A] | 16.5±2.4, n=35[A] | 16.6±2.2, n=44[A] | 16.4±1.6, n=23[A] | 16.7±2.2, n=42[A] | 17.3±2.0, n=25[A] | 16.3±2.2, n=36[A] | P=0.79 |
| CRLB (%) NAA | 3.0±0.4 | 3.1±0.3 | 3.1±0.3 | 3.0±0.3 | 3.0±0.2 | 3.0±0.0 | 3.0±0.2 | 3.0±0.3 | P=0.09 |
| CRLB (%) Myo-Inositol | 4.1±0.4 | 4.1±0.5 | 4.3±0.7 | 4.1±0.5 | 4.4±0.6 | 4.1±0.3 | 4.0±0.4 | 4.0±0.4 | P=0.13 |
| CRLB (%) Choline | 3.1±0.3 | 3.1±0.2 | 3.2±0.5 | 3.0±0.2 | 3.1±0.2 | 3.0±0.1 | 3.1±0.3 | 3.1±0.3 | P=0.52 |
| CRLB (%) PCr+Cr | 2.9±0.2 | 3.0±0.1 | 3.0±0.2 | 2.9±0.3 | 2.9±0.2 | 3.0±0.2 | 2.9±0.2 | 2.9±0.3 | P=0.71 |
| **Left thalamus** | | | | | | | | | |
| N | n=53 | n=49 | n=40 | n=45 | n=30 | n=41 | N=29 | N=37 | |
| CRLB (%) Gln | NA | NA | NA | NA | NA | NA | NA | NA | NA |
| CRLB (%) NAA | 5.1±0.9 | 5.0±0.9 | 5.3±1.7 | 5.0±0.9 | 4.9±0.9 | 4.9±0.8 | 5.1±1.0 | 4.9±1.1 | P=0.65 |
| CRLB (%) Myo-Inositol | 8.7±1.8 | 8.2±1.8 | 8.8±2.0 | 8.3±1.7 | 8.0±1.4 | 8.5±2.1 | 8.2±1.7 | 8.6±1.9 | P=0.82 |
| CRLB (%) Choline | 4.5±0.9 | 4.4±0.8 | 4.5±0.9 | 4.4±0.7 | 4.3±0.7 | 4.4±0.8 | 4.3±0.8 | 4.2±0.9 | P=0.72 |
| CRLB (%) PCr+Cr | 4.0±0.8 | 3.8±0.8 | 3.8±0.7 | 3.8±0.7 | 3.7±0.7 | 3.9±0.7 | 3.9±0.7 | 3.7±0.8 | P=0.65 |

Abbreviations: FEP: First episode psychosis; HC: Healthy controls; Cramér-Rao lower bound; NA: Not applicable; Glx: Glutamate+glutamine; NAA: N-acetyl aspartate; PCr+Cr: Total creatine (creatine+phosphocreatine). [A]: n is stated for the individual metabolite if data were excluded due to CRLB>20%. Glutamine data in left thalamus are not reported due to CRLB>20% in 85% of the baseline spectra.

Group differences in quality measures during the two years were tested with linear mixed models. The group*time interaction terms were insignificant for all CRLB values and were therefore removed from the statistical models.

**Statistics**

Changes in the relationship between glutamate levels in dACC and left thalamus over two years in FEP and HC were explored in the following linear mixed model:

$$Glu_{dACC} = \beta_0*Glu_{Thal} + \beta_1*group + \beta_2*sex + \beta_3*age + \beta_4*smoking + \beta_5*CRLB_{dACC} + \beta_6*FWHM_{dACC} + \beta_7*group*time*Glu_{Thal}$$

$Glu_{dACC}$: Glutamate levels in dorsal anterior cingulate cortex
$Glu_{Thal}$: Glutamate levels in left thalamus
$CRLB_{dACC}$: Cramér-Rao lower bound values for dACC
$FWHM_{dACC}$: Full width at half maximum for dACC

In case of a significant group*time* $Glu_{Thal}$ interaction, the following two similar linear mixed models were performed. First, a linear mixed model where the interaction-term in the model above was substituted with time* $Glu_{Thal}$ to evaluate, if the change over time was due to change in the relationship between $Glu_{dACC}$ and $Glu_{Thal}$ in the combined group of FEP and HC over the two years. Second, the interaction-term in the model above was substituted with a group* $Glu_{Thal}$ to evaluate, if the change was due to a different relationship between FEP and HC at all visits.

### Supplementary Results

*Mean levels of metabolites*

Mean levels of glutamatergic metabolites over the four visits are provided in Supplementary Table S4 for dACC and left thalamus.

**Supplementary Table S4: Mean levels of metabolites in dorsal anterior cingulate cortex and left thalamus**

| **Dorsal anterior cingulate cortex metabolite levels** | | | | | | | | |
|---|---|---|---|---|---|---|---|---|
| | **Baseline** | | **6 weeks** | | **6 months** | | **2 years** | |
| | **FEP** | **HCs** | **FEP** | **HCs** | **FEP** | **HCs** | **FEP** | **HCs** |
| N | N=54 | N=53 | N=42 | N=48 | N=31 | N=49 | N=33 | N=43 |
| Glutamate ± SD | 10.23±0.78 | 10.49±0.67 | 10.22±0.96 | 10.28±0.67 | 10.08±0.60 | 10.23±0.65 | 9.91±0.86 | 10.24±0.61 |
| Glx ± SD | 13.50±1.25 | 13.74±1.09 | 13.51±1.75 | 13.61±0.99 | 13.36±1.02 | 13.44±0.98 | 12.92±1.26 | 13.51±0.93 |
| Gray matter % ± SD | 69.4±3.0 | 69.2±2.3 | 69.6±3.5 | 69.0±2.3 | 69.4±2.9 | 69.7±2.7 | 69.3±4.1 | 70.6±3.6 |
| White matter % ± SD | 15.4±3.4 | 15.2±2.7 | 15.5±3.6 | 15.6±3.0 | 15.4±2.7 | 15.2±2.9 | 15.8±3.7 | 14.6±3.1 |
| **Left thalamic metabolite levels** | | | | | | | | |
| | **Baseline** | | **6 weeks** | | **6 months** | | **2 years** | |
| N | N=53 | N=49 | N=40 | N=45 | N=30 | N=41 | N=29 | N=37 |
| Glutamate ± SD | 6.84±0.83 | 6.74±0.81, n=48[A] | 6.80±0.77 | 6.84±0.73 | 6.65±0.78 | 7.10±0.96 | 6.73±0.90 | 6.85±0.84 |
| Glx ± SD | 9.62±1.29 | 9.75±1.33 | 9.65±1.51 | 9.76±1.16 | 9.57±1.35 | 9.90±1.37 | 9.47±1.20 | 9.44±1.14 |
| Gray matter % ± SD | 10.3±5.6 | 8.5±2.9 | 11.8±5.7 | 9.1±3.6 | 9.0±3.3 | 11.0±5.3 | 10.9±5.7 | 11.9±6.6 |
| White matter % ± SD | 89.5±5.7 | 91.3±2.9 | 88.0±5.9 | 90.7±3.7 | 90.8±3.3 | 88.9±5.4 | 88.9±5.9 | 87.9±6.6 |

Supplementary Table S4 shows mean metabolite levels and gray- and white matter fraction in dorsal anterior cingulate cortex and left thalamus in institutional units at all four visits in first-episode patients and healthy controls. [A]: n is stated for single metabolite levels if single subject data were excluded due to CRLB>20%. Abbreviations: ACC: Anterior cingulate cortex; FEP: First episode psychosis; HC: Healthy controls; SD: standard deviation; Glx: Glutamate+glutamine.

**Supplementary Table S5: Mean levels of other metabolites in dorsal anterior cingulate cortex and left thalamus**

| | **Baseline** | | **6 weeks** | | **6 months** | | **2 years** | |
|---|---|---|---|---|---|---|---|---|
| | **FEP** | **HCs** | **FEP** | **HCs** | **FEP** | **HCs** | **FEP** | **HCs** |
| **Dorsal anterior cingulate cortex** | | | | | | | | |
| N | N=54 | N=53 | N=42 | N=48 | N=31 | N=49 | N=33 | N=43 |
| Glutamine ± SD | 3.44±0.68, n=45[A] | 3.50±0.47, n=42[A] | 3.48±0.76, n=35[A] | 3.38±0.50, n=44[A] | 3.55±0.44, n=23[A] | 3.32±0.46, n=42[A] | 3.24±0.48, n=25[A] | 3.42±0.43, n=36[A] |
| NAA ± SD | 8.38±0.41 | 8.52±0.40 | 8.42±0.47 | 8.46±0.36 | 8.44±0.44 | 8.46±0.45 | 8.36±0.51 | 8.35±0.43 |
| Myo-Inositol ± SD | 5.66±0.46 | 5.76±0.44 | 5.62±0.44 | 5.76±0.47 | 5.63±0.35 | 5.74±0.42 | 5.67±0.62 | 5.68±0.41 |
| Choline ± SD | 1.90±0.20 | 1.95±0.16 | 1.89±0.21 | 1.96±0.17 | 1.92±0.13 | 1.96±0.17 | 1.88±0.21 | 1.96±0.19 |
| PCr+Cr ± SD | 6.82±0.31 | 6.80±0.36 | 6.79±0.36 | 6.74±0.33 | 6.85±0.35 | 6.71±0.42 | 6.68±0.41 | 6.69±0.32 |
| **Left thalamus** | | | | | | | | |
| N | N=53 | N=49 | N=40 | N=45 | N=30 | N=41 | N=29 | N=37 |
| Glutamine ± SD | NA | NA | NA | NA | NA | NA | NA | NA |
| NAA ± SD | 7.12±0.50 | 7.29±0.46 | 7.15±0.67 | 7.25±0.36 | 7.12±0.37 | 7.29±0.40 | 7.21±0.48 | 7.37±0.55 |
| Myo-Inositol ± SD | 3.46±0.52 | 3.59±0.46 | 3.47±0.46 | 3.61±0.53 | 3.56±0.34 | 3.49±0.49 | 3.64±0.56 | 3.35±0.54, N=36[A] |
| Choline ± SD | 1.66±0.19 | 1.70±0.15 | 1.71±0.17 | 1.69±0.14 | 1.70±0.17 | 1.70±0.13 | 1.71±0.16 | 1.73±0.17 |
| PCr+Cr ± SD | 5.47±0.48 | 5.53±0.29 | 5.71±0.38 | 5.49±0.28 | 5.64±0.31 | 5.51±0.27 | 5.57±0.41 | 5.61±0.37 |

Supplementary Table S5 shows mean metabolite levels in dorsal anterior cingulate cortex and left thalamus in institutional units at all four visits in first-episode patients and healthy controls. [A]: n is stated for single metabolite levels if single subject data were excluded due to CRLB>20%.
Abbreviations: ACC: Anterior cingulate cortex; FEP: First episode psychosis; HC: Healthy controls; SD: standard deviation; Glx: Glutamate+glutamine; NAA: N-acetyl aspartate; PCr+Cr: Total creatine (creatine+phosphocreatine).

**Supplementary Table S6: Differences in the trajectory or main effects of metabolites over two years in first-episode patients and healthy controls**

| **Metabolite levels** | **Group*time** | **Post hoc tests** | **Group** | **Time** | **Sex** | **Smoking (No of cigarettes)** | **Age of participants** |
|---|---|---|---|---|---|---|---|
| **Dorsal Anterior Cingulate Cortex** | | | | | | | |
| Glutamine | P=0.18 | NA | P=0.84 | P=0.12 | **P=0.01** Lower in females | P=0.44 | P=0.42 |
| NAA | P=0.53 | NA | P=0.48 | **P=0.02** Decrease with time | P=0.14 | P=0.22 | P=0.12 |
| Myo-Inositol | P=0.88 | NA | P=0.16 | P=0.11 | P=0.46 | P=0.12 | **P=0.03** Increase with higher age |
| Choline | P=0.11 | NA | P=0.27 | P=0.25 | **P=0.001** Lower in females | P=0.63 | **P=0.002** Increase with higher age |
| PCr+Cr | P=0.24 | NA | P=0.76 | **P=0.01** Decrease with time | P=0.09 | **P=0.05** Higher in smokers | P=0.09 |
| Gray matter[A] % | P=0.27 | NA | P=0.87 | P=0.08 | **P=0.02** Lower in females | P=0.19 | **P=0.02** Decrease with higher age |
| White matter[A] % | P=0.45 | NA | P=0.53 | P=0.43 | P=0.13 | P=0.60 | **P=0.02** Increase with higher age |

| | Group*time | Post hoc tests | Group | Time | Sex | Smoking (No of cigarettes) | Age of participants |
|---|---|---|---|---|---|---|---|
| **Left thalamus** | | | | | | | |
| Glutamine | NA | NA | NA | NA | NA | NA | NA |
| NAA | P=0.78 | NA | **P=0.04** FEP<HC | P=0.51 | P=0.89 | P=0.95 | P=0.08 |
| Myo-Inositol | P=0.05 | Baseline: p=0.35<br>6 weeks: p=0.35<br>6 months: p=0.75<br>2 years: **p=0.02** (FEP>HC) | NA | NA | P=0.15 | P=0.46 | **P=0.001** Increase with higher age |
| Choline | P=0.34 | NA | P=0.79 | P=0.32 | **P<0.001** Lower in females | P=0.78 | P=0.10 |
| PCr+Cr | **P=0.003** | Baseline: p=0.37<br>6 weeks: **p=0.01** (FEP>HC)<br>6 months: p=0.08 (FEP>HC)<br>2 years: p=0.49 | NA | NA | P=0.79 | P=0.84 | **P=0.01** Increase with higher age |
| Gray matter[A] % | **P=0.03** | Baseline: **p=0.02** (FEP>HC)<br>6 weeks: **p=0.03** (FEP>HC)<br>6 months: p=0.14<br>2 years: p=0.31 | NA | NA | **P=0.005** Higher in females | P=0.63 | P=0.91 |
| White matter[A] % | **P=0.03** | Baseline: **p=0.03** (FEP<HC)<br>6 weeks: **p=0.02** (FEP<HC)<br>6 months: p=0.14<br>2 years: p=0.29 | NA | NA | **P=0.004** Lower in females | P=0.65 | P=0.90 |

Table S6 shows a different trajectory of metabolites (group*time) followed by post-hoc tests for individual visits in case of significant group*time interaction or main effects of group and time in case of an insignificant interaction. Main effects for the nuisance variables sex, number of cigarettes and age of participants at study inclusion are shown as well. [A]: Gray and white matter % denotes the percentage in the spectroscopic voxel.

Abbreviations: No: Number; Glx: Glutamate+glutamine; NAA: N-acetyl aspartate; PCr+Cr: Total creatine (creatine+phosphocreatine); FEP: First episode psychosis; HC: Healthy controls

**Supplementary Figure S2**

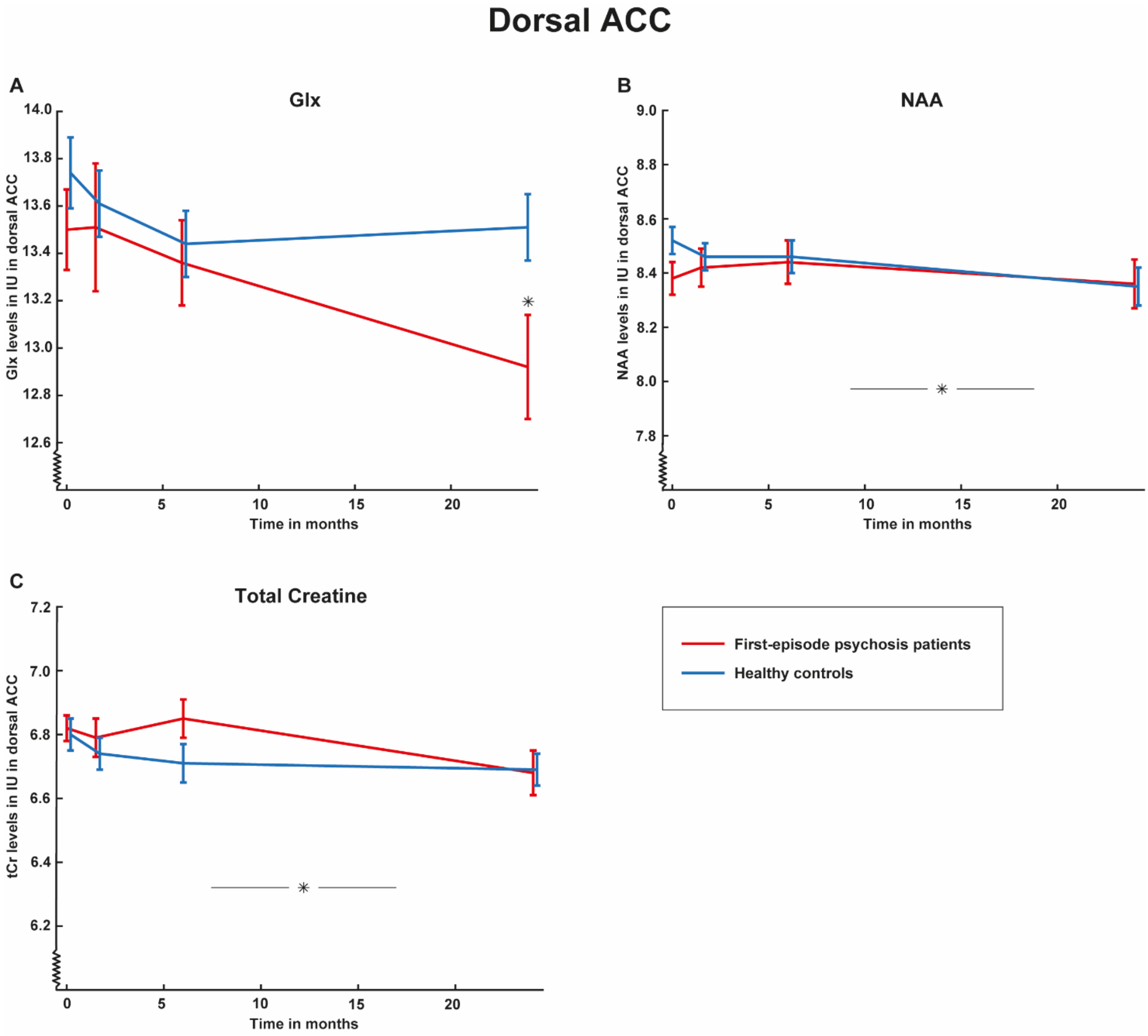


Supplementary Figure S2 shows mean metabolite levels and standard errors for Glx (glutamate+glutamine) (A), N-acetylaspartate (NAA) (B), and total creatine (C) in dorsal anterior cingulate cortex over two years in the initially antipsychotic-naïve first-episode psychosis patients (FEP) (red lines) compared with healthy controls (HC) (blue lines). Participants were assessed at baseline (0 months), after six weeks (1.5 months), six months, and two years (24 months). A: The trajectory of Glx levels were different in FEP compared with HC (group*time: $p=0.04$) due to lower glx levels in FEP after two years ($p=0.02$). B: NAA levels in dACC decreased significantly over the two years in the combined group of FEP and HC (main effect of time: $p=0.02$). C: Total creatine decreased significantly over the two years in the combined group of FEP and HC (main effect of time: $p=0.01$). Vertical bars represent main effect of time.
*: $p<0.05$.

**Supplementary Figure S3**

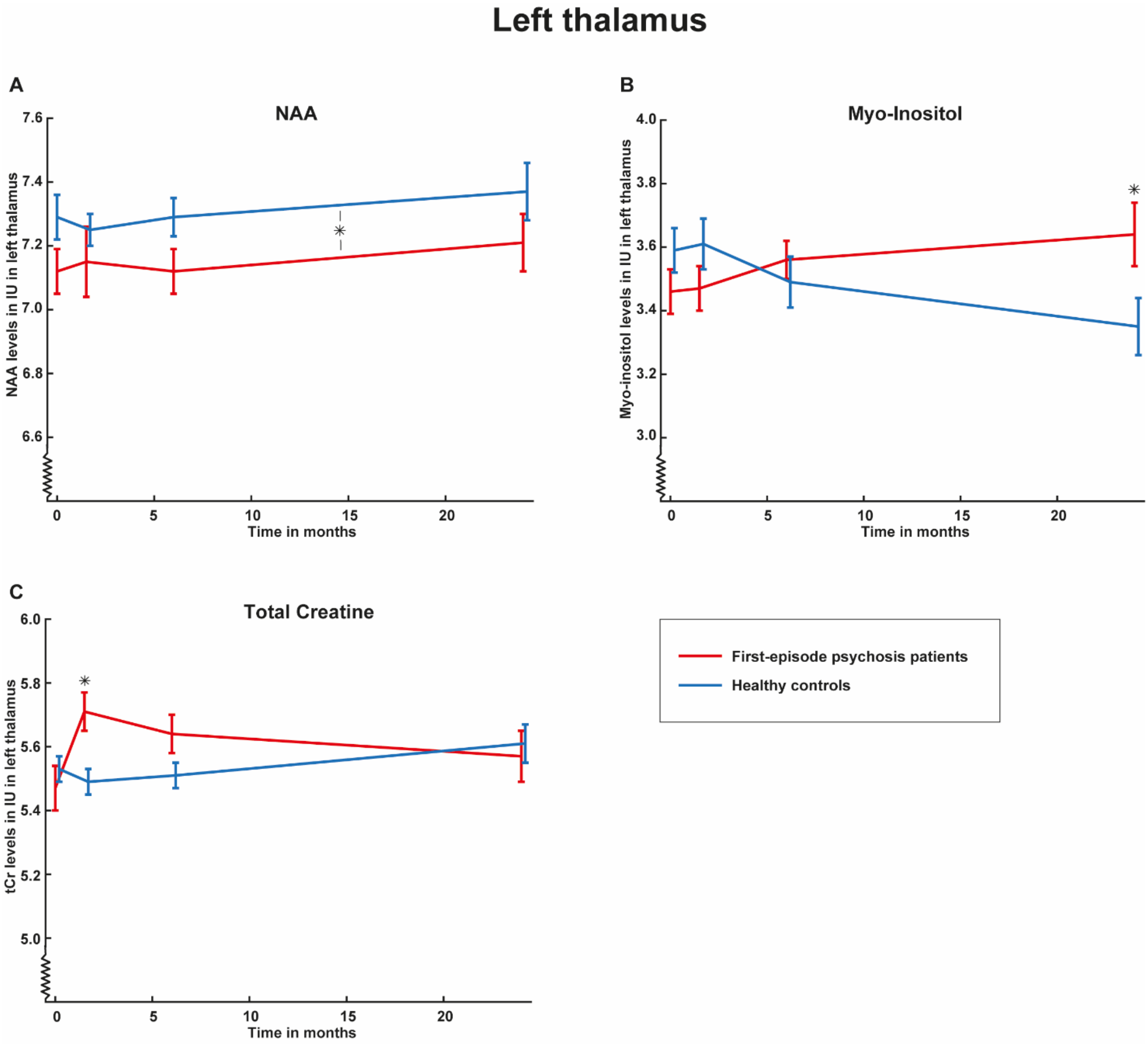


Supplementary Figure S3 shows mean metabolite levels and standard errors for N-acetylaspartate (NAA) (A), Myo-inositol (B), and total creatine (C) in left thalamus over two years in the initially antipsychotic-naïve first-episode psychosis patients (FEP) (red lines) compared with healthy controls (HC) (blue lines). Participants were assessed at baseline (0 months), after six weeks (1.5 months), six months, and two years (24 months). A: NAA levels in left thalamus were significantly lower in FEP at all assessments (main effect of group: p=0.04). B: Myo-inositol levels in left thalamus had a borderline significant different trajectory in FEP compared with HC (group*time: p=0.05) due to higher levels in FEP after two years (p=0.02). C: Total Creatine levels in left thalamus had a significantly different trajectory in FEP compared with HC (group*time: p=0.003) due to higher levels in FEP after 6 weeks of treatment (p=0.01). The horisontal bar represents main effect of group.
*: p<0.05.

*Other metabolites over two years in patients and healthy controls analyzed separately*

*Dorsal ACC*: When analyzing FEP only, there was a significant main effect of time for tCr (p=0.024) and borderline significant for choline (p=0.05) due to a decrease over two years, but no changes over time for glutamine (p=0.54), NAA (p=0.30), or myo-inositol (p=0.33). For HC, there were no significant changes over time for glutamine (p=0.14), NAA (p=0.09), myo-inositol (p=0.47), choline (p=0.75), or tCr (p=0.20).

*Left thalamus*: When analyzing FEP only, there was a significant main effect of time for tCr (p=0.014) due to an increase over time, but no significant changes over time for glx (p=0.94), NAA (p=0.12), myo-inositol (p=0.31), or choline (p=0.13). For HC, the main effect of time was insignificant for glx, NAA, myo-inositol, choline, and tCr (p=0.13-0.93).

*The association between glutamatergic metabolites in dACC and left thalamus over two years*

Exploratory analyses revealed that the association between glutamate levels in dACC and left thalamus changed over the two years ($Glu_{Thal}$*group*time: p=0.01) due to a change in the relationship between $Glu_{dACC}$ and $Glu_{Thal}$ in the combined group of FEP and HC over two years (time* $Glu_{Thal}$: p=0.016) and not a different relationship between $Glu_{dACC}$ and $Glu_{Thal}$ in FEP and HC at all visits (group* $Glu_{Thal}$: p=0.98).

Post hoc tests of the association between $Glu_{dACC}$ and $Glu_{Thal}$ at the four visits in FEP and HC indicated that this was due to a positive association between glutamate levels in dACC and left thalamus at 6 weeks (p<0.001, β=0.35, Supplementary Figure S4B), but a non-significant trend for a negative association after two years (p=0.14, β=-0.14, Supplementary Figure S4D). There were not significant associations at baseline (p=0.21, β=0.10, Supplementary Figure S4A) or after 6 months (p=0.97, β=0.003, Supplementary Figure S4C).

### Supplementary Figure S4

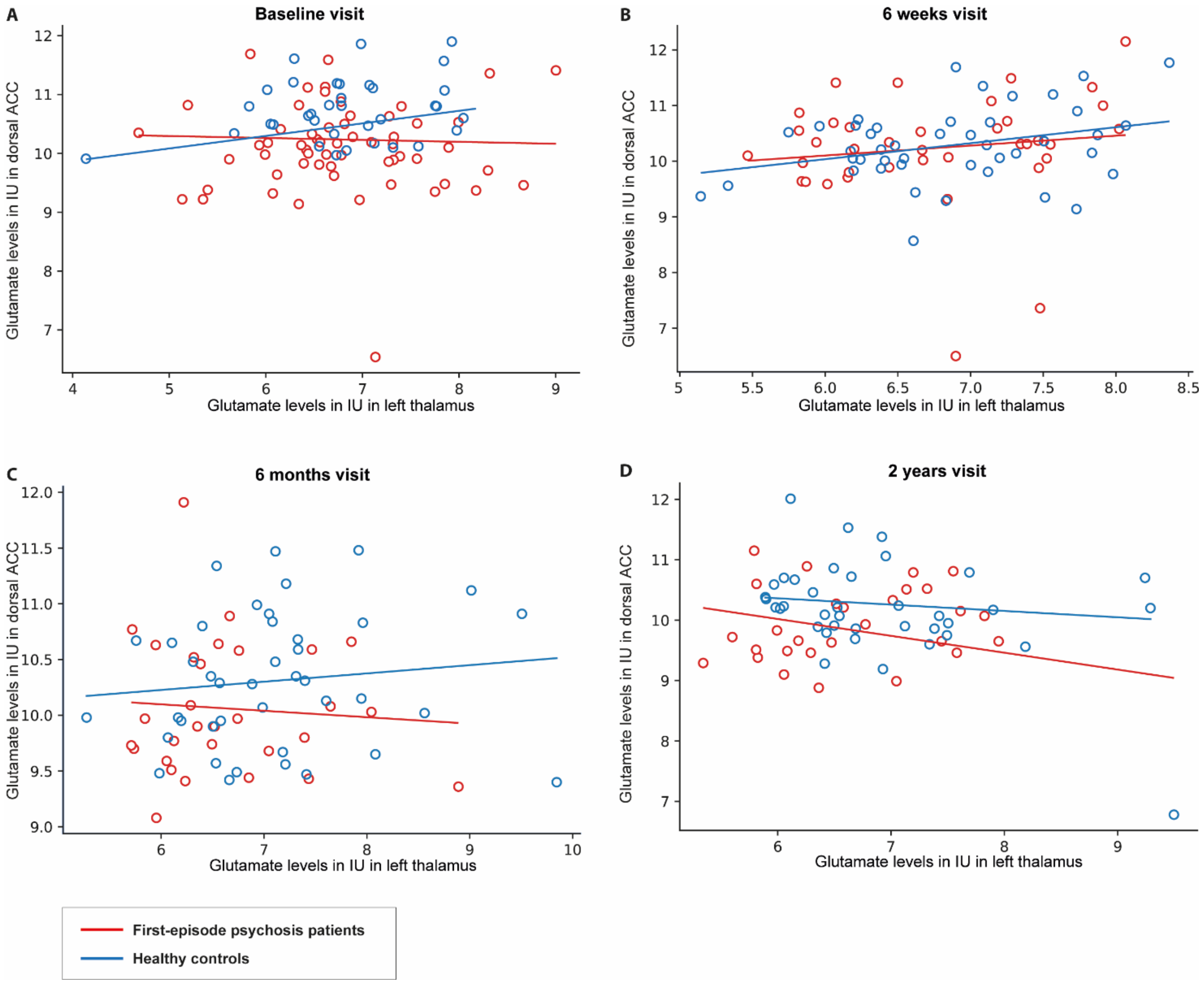


Supplementary Figure S4 illustrates the relationship between glutamate levels in dorsal anterior cingulate cortex ($Glu_{dACC}$) and left thalamus ($Glu_{Thal}$) in initially antipsychotic-naïve patients with first-episode psychosis (FEP) in (red circles) and healthy controls (blue circles) at baseline (A), after 6 weeks (B), 6 months (C), and 2 years (D). There was a significant change in the association between $Glu_{dACC}$ and $Glu_{Thal}$ over the two years ($Glu_{Thal}$*group*time: $p=0.01$) due to a change in the association between $Glu_{dACC}$ and $Glu_{Thal}$ in the combined group of FEP and HC (glutamate thalamus*time: $p=0.016$). Post hoc tests indicated that this may be due to a positive association between $Glu_{dACC}$ and $Glu_{Thal}$ after 6 weeks (B: $p<0.001$, $\beta=0.35$) but a non-significant trend for a negative association after 2 years (D: $p=0.14$, $\beta=-0.14$).

*Drop-out analyses*

Drop-out analyses comparing baseline PANSS positive and negative sub-scores and glutamate levels in patients completing and not completing the two-year follow-up visit revealed no significant differences as summarized in Supplementary Table S7.

**Supplementary Table S7: Drop-out analyses**

| | **Patients completing two-year follow-up visit (N)** | **Patients not completing two-year follow-up visit (N)** | **Statistics** |
|---|---|---|---|
| **DACC mean baseline glutamate levels IU ± SD** | 10.19±0.92 (33) | 10.28±0.51 (21) | F=0.21, p=0.65 |
| **Left thalamic mean baseline glutamate levels IU ± SD** | 6.89 ± 0.99 (32) | 6.77 ± 0.52 (21) | F=1.34, p=0.25 |
| **PANSS positive subscore ± SD at baseline** | 18.7 ± 4.2 (35) | 18.8 ± 4.6 (22) | F=0.00, p=0.97 |
| **PANSS negative subscore ± SD at baseline** | 20.4 ± 5.3 (35) | 19.0 ± 5.3 (22) | F=0.81, p=0.37 |

Supplementary Table S4 shows mean levels of glutamate as well as mean PANSS positive and negative sub-scores in first-episode patients with psychosis completing or not completing the two year follow-up visit.
Abreviations: dACC: dorsal Anterior Cingulate Cortex; IU: Institutional Units; SD: Standard deviation. PANSS: Positive and negative syndrome scale.
